# Synaptic density deficits in patients with a psychotic disorder and unaffected siblings

**DOI:** 10.1101/2025.06.05.25328865

**Authors:** Monique van der Weijden-Germann, Jesca E. de Jager, Jasper O. Nuninga, Erik F.J. de Vries, Chris W.J. van der Weijden, Iris E.C. Sommer

## Abstract

**Background:** Schizophrenia spectrum disorders (SSD) are associated with cognitive dysfunction, which has been linked to excessive synaptic pruning. To disentangle SSDs genetic vulnerability from secondary effects, we investigated synaptic density in individuals with SSD, unaffected siblings of individuals with SSD, and healthy controls (HC).

**Methods:** We recruited 24 SSD individuals, 25 unaffected siblings, and 26 HC. Participants underwent cognitive assessments using the Brief Assessment of Cognition in Schizophrenia and symptom severity in individuals with SSD was assessed with the Positive and Negative Syndrome Scale. Synaptic density was measured using [^11^C]UCB-J PET., with binding potential (BP_ND_) calculated across multiple *a priori* chosen regions of interest (ROIs). Group differences in BPND and associations with cognition and symptoms were assessed using multiple linear regression and correlation.

**Results:** SSD individuals had significant lower cognitive functioning and reduced [^11^C]UCB-J BP_ND_ in frontal, temporal, and limbic regions, including the hippocampus, amygdala, superior temporal gyrus, insula, and inferior frontal gyrus, compared to HC. Siblings exhibited preserved cognitive performance but displayed significant BP_ND_ reductions in overlapping regions as SSD individuals, including the hippocampus, amygdala, and superior temporal gyrus, relative to HC. Across all participants, verbal memory performance correlated with BP_ND_ in the left superior frontal gyrus, precentral gyrus, and right middle frontal gyrus. Motor speed was associated with BP_ND_ in the cerebellum.

**Conclusion:** Despite deficits in synaptic density, siblings maintained intact cognitive performance, suggesting compensatory mechanisms. Our findings support a relationship between synaptic pathology, genetic liability, and cognitive resilience, which may inform targeted interventions for SSD.

## 1. Introduction

Schizophrenia spectrum disorders (SSD) are estimated to affect ~24 million people worldwide [1] with a lifetime prevalence of 1.25% [2], placing a significant burden on global health. SSD are characterized by positive symptoms, such as hallucinations and delusions, negative symptoms, such as apathy, and cognitive dysfunction across domains [3]. Antipsychotic medication effectively mitigates positive symptoms in the majority of individuals with SSD, but effects on cognition are absent [4]. Neurocognitive functioning in persons with SSD is relatively stable over time and was consistently shown to correlate negatively with daily functioning, with the majority of patients failing to achieve long-term partnership and financial independence [5, 6]. Consequently, neurocognitive functioning can be an important marker of social and functional prognosis [7]. Yet, neuropsychological test results in schizophrenia do not correspond very well with real-life functioning [8], as test performance can be affected by decreased motivation. As motivational syndrome is a fundamental aspect of the negative symptoms of schizophrenia, more reliable reflections of cognitive capacity are needed. Currently, few treatments exist that can curtail cognitive dysfunction [9] and several drugs that may be effective are under development [10]. For the evaluation of these drugs and the further development of both pharmacological and non-pharmacological pro-cognitive interventions, thorough understanding of the underlying pathophysiology of cognitive impairments in SSD is crucial. Although it is now widely accepted that cognitive dysfunction has a multifaceted etiology [11, 12], many questions regarding its etiology remain unanswered.

The synaptic pruning hypothesis, originally proposed by Irwin Feinberg (1982), is among the earliest and still influential theories regarding the pathophysiology of cognitive dysfunction in SSD. Feinberg describes a crucial role of excessive pruning of synapses leading to decreased and less effective connectivity [13]. First psychotic symptoms in individuals with SSD tend to become manifest when brain areas involved in higher cognitive functions are subjected to excessive pruning [14]. In SSD, the pruning process may not only be limited to infrequently used synapses but may also include frequently used connections, giving rise to insufficient connectivity [13]. Indeed, connectivity deficits have been demonstrated in individuals with SSD, both at the macroscopic and microscopic level, which correlate with cognitive dysfunction [15].

Studies using induced pluripotent stem cells and genome-wise association studies, have confirmed and extended the pruning hypothesis, assigning a crucial role to genetically induced complement-mediated abnormal synaptic pruning by microglia [16–19]. Currently, the precise molecular mechanisms of increased pruning in SSD, its dependence of genetic vulnerability and the role of environmental factors, like medication use, remain unclear.

Positron emission tomography (PET) with the radioligand [^11^C]UCB-J has opened the way to noninvasive in-vivo assessment of synaptic density [20]. [^11^C]UCB-J binds to synaptic vesicle glycoproteins 2A (SV2A), an integral 12-transmembrane domain glycoprotein expressed in pre-synaptic vesicles throughout the brain. Studies showed high correlations between SV2A expression and density of presynaptic terminals [21]. Based on these correlations, [^11^C]UCB-J binding may be considered as a proxy for synaptic density. The first studies applying this radiotracer in individuals with SSD demonstrated reductions in tracer binding in the frontal and anterior cingulate cortices (ACC) [22], hippocampus [23], putamen, and superior temporal gyrus [24], as compared to healthy controls (HC). Furthermore, [^11^C]UCB-J binding expressed as non-displaceable binding potential (BP_ND_), was positively associated with overall cognitive ability, speed of detection, social cognition and negatively associated with symptom severity [23, 24].

Radhakrishnan et al. (2021) did not find a correlation between synaptic density and cumulative dose of antipsychotic in 13 participants [23], which argues against a secondary effect, but could also be due to insufficient statistical power. A recent PET study using the same tracer included both first episode patients and people with clinical high risk. Compared to healthy controls, both groups had lower synaptic density, which was related to more severe negative symptoms, but also to cannabis use [25].

Siblings of individuals with SSD share part of the genetic background for schizophrenia [26]. If genetic vulnerability plays an essential role in processes shaping synaptic density, aberrant synaptic density should also be detectable in unaffected siblings of individuals with SSD. The current study set out to investigate synaptic density and its relation to cognitive functioning in a fairly large sample of individuals with SSD, unaffected siblings, and healthy individuals matched for age and sex. The primary aim was to investigate the extent to which abnormal synaptic density, as measured with [^11^C]UCB-J, can also be observed in siblings of patients with SSD, reflecting the genetic vulnerability for SSD. As a secondary aim, we investigated whether such alterations in synaptic density affect cognitive functioning of individuals with SSD and their siblings. Finally, we investigated potential relationships of synaptic density with symptom severity at time of investigation.

## 2. Methods and Materials

The present study protocol was approved by the local medical and ethical committee of the University Medical Centre Groningen (UMCG) (METc number: NL7110004219, Protocol ID: 201900529, Trial registration number: NL-OMON27759). All study related procedures were conducted in accordance with the Declaration of Helsinki (64^th^ WMA general assembly; October 2013). Before participation, all study subjects provided written informed consent.

### 2.1. Participants

In total, 24 patients with SSD, 25 unaffected siblings of patients with SSD, and 26 healthy controls were included for the present study. Participants were recruited via social media, word of mouth, newsletter advertisement, and patient associations. Individuals were included if they spoke Dutch as native language, were between 26 and 65 years old and able to understand the study and provide written informed consent for participation. For the patient group, participants needed to be diagnosed with schizophrenia, schizoaffective disorder, or schizophreniform disorder (295.x) without the occurrence of dangerous or harmful behavior within 3 months prior to inclusion. The diagnosis, duration, and severity of disease was confirmed by the Comprehensive Assessment of Symptoms and History (CASH) interview [27]. For individuals in the sibling group, the inclusion criteria of having a sibling with a diagnosis of schizophrenia, schizoaffective disorder, or schizophreniform disorder (295.x) was applied. In addition, siblings had to be free of any psychiatric diagnoses themselves. Siblings were not necessarily related to participants of the patient group. Finally, individuals from the healthy control group were required to be free of any psychiatric or neurological disease and to not have any first- or second-degree relatives with one of the disorders as specified for the patient group.

### 2.2. Cognitive and Clinical assessments

Neurocognitive functioning was assessed with the Dutch version of the Brief Assessment of Cognition in Schizophrenia (BACS; detailed description in Supplementary material), including the following 6 sub-tests [28]:

1. List learning: verbal memory.
2. Digit sequencing task: working memory.
3. Token motor task: motor speed.
4. Category instances and controlled oral association test: verbal fluency.
5. Symbol coding: attentional and speed of information processing.
6. Tower of London: executive functioning.

In addition, the Stroop task [29], measuring executive functioning and verbal inhibition, and the Symbol Search task, originating from the WAIS-IV [30] and measuring information processing speed, were administered. To ensure the highest quality of data, administration of all tests and interviews was done by comprehensively trained study personnel.

Raw performance scores on all subtests of the BACS were converted into age and gender adjusted standardized z-scores in accordance with procedures described by [31]. By averaging all standardized subtest z-scores, the composite BACS score, reflecting overall cognitive performance, was computed. For the Stroop task, the interference factor was calculated by subtracting the reaction time (RT) on card C by the RT on card B individually per subject [32]. Use of antipsychotic medication at the time of study assessments was documented based on questionnaires filled in by the patient. Reported dose of antipsychotic medication usage (mg/day) was transformed into an antipsychotic dose equivalent (i.e.: olanzapine equivalent) [33]. Finally, the highest achieved educational level as derived from the CASH was converted into quantitative years of education (YOE, Supplementary Table S1). Symptom severity at time of scanning was assessed by consensus score of two trained investigators using the Positive and Negative Syndrome Scale (PANSS) [34]. For analyses we used positive, negative, general, and total symptom severity.

### 2.3. Magnetic Resonance Imaging (MRI)

For registration of the PET-data, a structural MRI-scan was acquired on a 3.0 Tesla Siemens scanner (Siemens Magnetom Prisma) equipped with a 64-channel head coil (Siemens, Erlangen, Germany). The MRI acquisition protocol entailed a sagittal 3D T_1_w MPRAGE-image with the following parameters: repetition time: 2300 ms, time to echo: 2.98 ms, inversion time: 900 ms, flip angle: 9°, voxel size: 1 mm isotropic.

### 2.4. Positron Emission Tomography

To ensure no pregnant women were included, female participants were tested for pregnancy prior to the PET-scan. PET-scans were performed on a Siemens Biograph mCT Vision PET/CT scanner and scan data were acquired in list mode format. Prior to PET acquisition, a low-dose CT-scan was performed for attenuation correction. Participants were intravenously injected with [^11^C]UCB-J at the start of a 60-minute dynamic PET acquisition. Individual doses of [^11^C]UCB-J were prepared on site and in accordance with GMP quality assurance criteria. PET-data were corrected for randoms scatter, dead-time, radioactive decay, and attenuation, and reconstructed in 25 frames (11 × 10, 4 × 60, 2 × 120, 2 × 180, 5 × 300, and 1 × 600s) with a voxel size of 0.9 mm isotropic.

#### 2.4.1. PET-data preprocessing

The preprocessing and analysis of PET images were conducted using the PMOD Neuro Tool, version 4.105 (PMOD Technologies LLC, Zurich, Switzerland). First, motion of the participant during PET acquisition was evaluated by assessment of the alignment of each individual frame of the PET scan and with the low-dose CT in the axial, coronal, and sagittal direction. If PET motion was more than 4 mm (i.e.: the spatial resolution of the Siemens biograph mCT Vision) [35] in one of the three directions, the corresponding PET images were subjected to motion correction. If motion was more than 2 times the PET scanner resolution (i.e.: >8 mm) in one of the three directions, the PET-scan was reconstructed again with a priori CT alignment to the images. Reconstructed PET images were co-registered to the T_1_w MRI, followed by segmentation of the PET images into individual brain regions using the Hammers atlas, to which a region of interest (ROI) for centrum semiovale was added. The centrum semiovale was manually delineated in MNI space by an expert neuro-imaging scientist and the accuracy of the delineation was confirmed by an experienced neuro-radiologist. The centrum semiovale was chosen as the reference region based on previous studies reporting lowest [^11^C]UCB-J radioactivity concentration in subcortical white matter regions, such as the centrum semiovale [36, 37]. Time-activity-curves (TACs) were generated for 48 ROIs (i.e., 24 brain regions, left and right hemisphere separately).

#### 2.4.2. Image analysis

Simplified Reference Tissue Model 2 (SRTM2) has previously been determined as the optimal quantification method for [^11^C]UCB-J PET [38]. Therefore, the present study applied parametric kinetic modelling to generate SRTM2 BP_ND_ images. This was performed as follows: first SRTM was employed according to previously reported optimal settings [38] with the centrum semiovale as a reference region to obtain k_2_’ images. The whole brain median SRTM k_2_’ was subsequently used as k_2_’ in SRTM2 with centrum semiovale as a reference region. Finally, the BP_ND_ per ROI was extracted from the SRTM2 BP_ND_ images.

### 2.5. Statistical analysis

Preprocessing of cognitive and demographic data as well as subsequent statistical analyses were performed in RStudio Version 4.1.2 (RStudio Team (2023), RStudio, Inc., Boston, MA (http://www.rstudio.com/). Cognitive performance differences between the groups were assessed with multiple linear regression (MLR) analyses using age and years of education as covariates. If Shapiro-Wilk normality tests and Q-Q-plots indicated non-normally distributed data, a data-transformation was applied using the R-package “bestNormalize” [39, 40]. After the application of the data-transformation, all assumptions were subsequently assessed again.

To assess group differences in [^11^C]UCB-J BP_ND_, we first performed MLR analyses with BP_ND_ in a set of *a priori* selected ROIs (see Supplementary material) as outcome variables and age and years of education as covariates. Next, we performed 6 different MLR analyses each with a distinct cognitive domain (i.e.: attention and speed of information processing, executive functioning, motor speed, verbal fluency, verbal memory, working memory) as outcome variable. For each cognitive domain, we used a unique set of *a priori* specified ROIs based on neuronal circuits involved with these tasks (Supplementary material, “*A priori ROI selection*”) as independent variable and age and years of education as covariates. All MLR analyses were preceded by the assessment of the statistical assumptions of linearity, homoscedasticity, (multivariate) normality, and independence of residuals. All follow-up comparison analyses were adjusted for multiple comparisons using the Benjamin Hochberg false-discovery rate (FDR) adjustment. That is, reported p-values are adjusted for multiple comparisons using an FDR correction method and were considered to be significant at an FDR-corrected p-value of ≤ 0.05.

We tested whether there were significant correlations between [^11^C]UCB-J BP_ND_ and PANSS positive, negative, general, and total scores in the individuals with SSD using Pearson’s product-moment correlation coefficients for normally distributed data and Spearman’s rank correlations for non-normally distributed data. The correlations between [^11^C]UCB-J BP_ND_ and cognitive outcomes and symptomatology were assessed in a set of *a priori* selected ROIs depending on the certain (cognitive) domain (see Supplementary material). The selection of ROIs for every cognitive domain as well as for every PANSS domain was based on previous literature (Supplementary material “*A priori ROI selection*”). FDR-corrected p-values of < 0.05 were considered significant. Correlations higher than r > 0.5 were considered as strong correlations, while correlations between r = 0.3 – 0.5 were considered as moderate and those smaller than r ≤ 0.3 were considered as weak correlations [41]. Finally, to exclude the possibility of differences in tracer uptake in the reference region between the groups, we performed an ANOVA assessing whether there are significant differences in [^11^C]UCB-J BP_ND_ in the centrum semiovale between the groups.

## 3. Results

### 3.1. Demographics

Table 1 displays the demographic characteristics of the full study sample and separately for participants with SSD (*N* = 24), unaffected siblings of patients (SIB, *N* = 25), and healthy controls (HC, *N* = 26). Even though the SSD and HC groups contained a greater proportion of male than female subjects as compared to the SIB group, there was no statistically significant difference in gender between the three groups (*X*^*2*^ = 4.61 *p* = 0.09). The groups were not statistically different with respect to age (*H* = 5.8, *p* = 0.055) or years of education (*H* = 4.9, *p* = 0.08). The groups did not show significant differences in [^11^C]UCB-J BP_ND_ in the centrum semiovale.

**Table 1.**
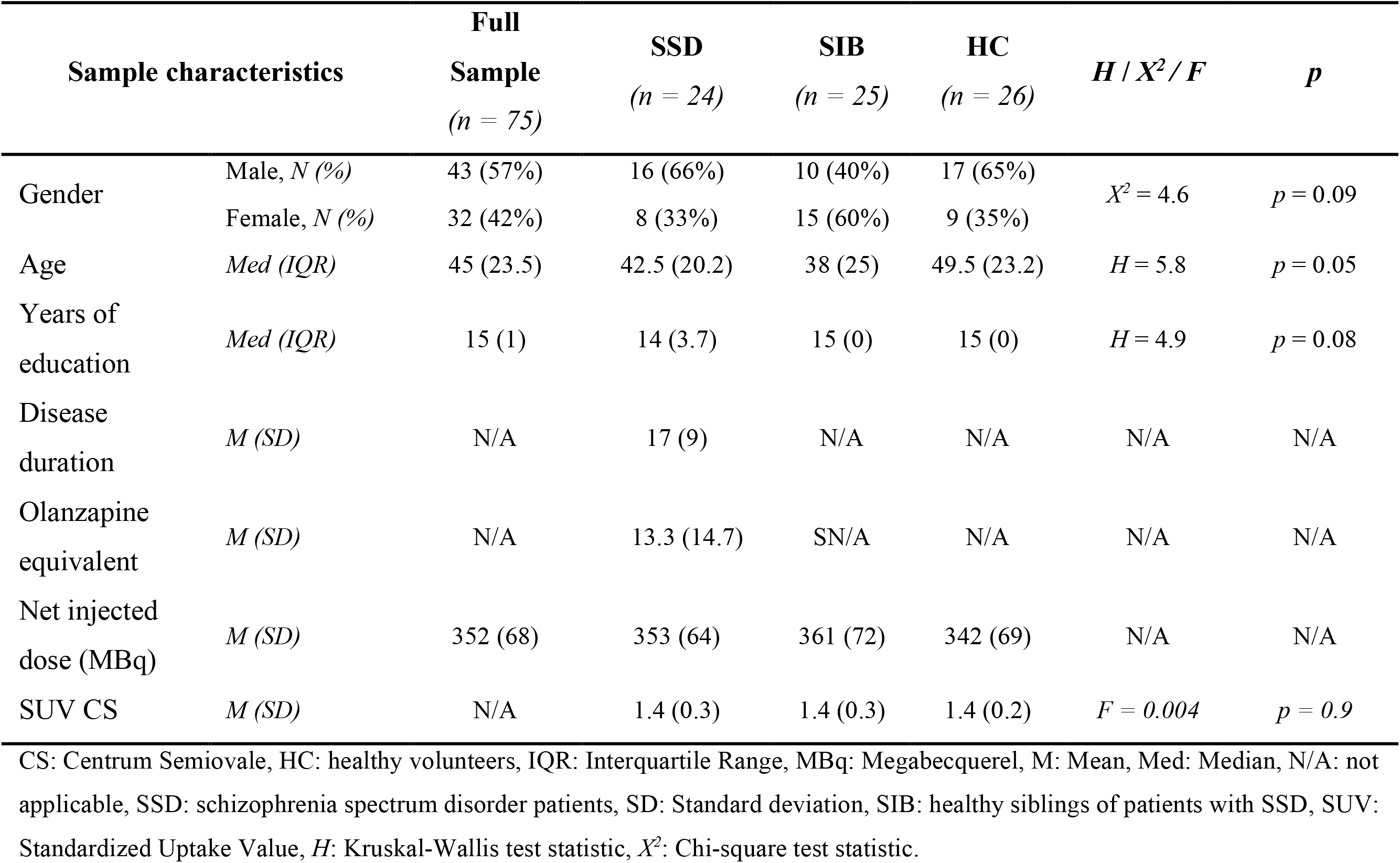
Demographic characteristics of schizophrenia spectrum disorder (SSD), unaffected siblings (SIB), and healthy control (HC) participants.

### 3.2. Group differences in Cognitive performance

Table 2 displays differences in cognitive performance between the groups for overall cognitive ability (i.e., BACS-composite score) as well as the 6 cognitive subdomains assessed by the BACS (Table 2; Supplementary material, figure S1). A significant effect of group was present in overall cognitive ability, verbal memory, verbal fluency, and attention and information processing speed. Individuals with SSD displayed significant impairment in overall cognitive ability compared to both HC *(p* = 0.007) and when compared to SIB (*p* = 0.015). Individuals with SSD performed significantly worse than SIB on verbal memory (*p* = 0.001) as well as verbal fluency (*p* = 0.006). On attention and information processing speed tasks, individuals with SSD performed significantly worse than HC (*p* = 0.026). Finally, for the two additional tests next to the BACS, no significant group differences were found for the Stroop task, while individuals with SSD performed significantly worse on the symbol search task as compared to their unaffected siblings (*p* = 0.005) and HC (*p* = 0.024). There were no significant differences in cognitive performance between healthy controls and siblings of individuals with SSD.

**Table 2.** Cognitive and clinical characteristics of schizophrenia spectrum disorder (SSD), unaffected siblings (SIB), and healthy control (HC) participants.

| BACS, z-scores |  | SSD<br>(n = 24) | SIB<br>(n = 25) | HC<br>(n = 26) | F / H | df | p | Post-hoc |
| --- | --- | --- | --- | --- | --- | --- | --- | --- |
| Composite score | M (SD) | -0.04 (0.07) | 0.01 (0.06) | 0.01 (0.05) | F=8.3 | 3 | p < 0.001*** | a, b |
| Verbal memory | M (SD) | -0.4 (1.4) | 0.5 (0.7) | 0.1 (0.9) | F=6.1 | 3 | p < 0.001*** | a |
| Working memory | Med (IQR) | -0.6 (2.0) | 0.3 (1.1) | 0.3 (1.1) | F=2.8 | 3 | p = 0.04 | --- |
| Motor speed | M (SD) | -0.2 (1.1) | 0.3 (0.9) | -0.09 (1.0) | F=1.4 | 3 | p = 0.2 | --- |
| Verbal fluency | M (SD) | -0.5 (1.1) | 0.2 (0.9) | 0.06 (0.9) | F=8.2 | 3 | p < 0.001*** | a |
| Attention & IPS | M (SD) | -0.5 (0.7) | -0.1 (0.9) | 0.1 (0.9) | F=4.2 | 3 | p < 0.01** | b |
| Executive functioning | Med (IQR) | 0.1 (0.7) | 0.1 (0.9) | 0.4 (0.6) | F=0.4 | 3 | p = 0.7 | --- |
| **Stroop task** |  |  |  |  |  |  |  |  |
| Interference factor | Med (IQR) | 27.6 (13.7) | 25.5 (12.4) | 25.3 (10.7) | F=3 | 3 | p = 0.03 | --- |
| **Symbol Search task** |  |  |  |  |  |  |  |  |
| Total score | M (SD) | 29.8 (7.0) | 36.2 (7.4) | 34.4 (6.3) | F=16.8 | 3 | p < 0.001*** | a, b |
| **PANSS** |  |  |  |  |  |  |  |  |
| Positive scale | Med (IQR) | 14 (9.2) | N/A | N/A | --- | --- | --- | --- |
| Negative scale | Med (IQR) | 11 (7.2) | N/A | N/A | --- | --- | --- | --- |
| General scale | Med (IQR) | 27.5 (14.5) | N/A | N/A | --- | --- | --- | --- |
| Total | Med (IQR) | 55.5 (25.5) | N/A | N/A | --- | --- | --- | --- |

a: SSD differ significantly from SIB, b: SSD differ significantly from HC, F: ANOVA test statistic, H: Kruskal-Wallis test statistic, HC: healthy volunteers, IQR: Interquartile Range, M: Mean, Med: Median, N/A: not applicable, PANSS: Positive And Negative Syndrome Scale, SSD: Schizophrenia Spectrum Disorder patients, SD: Standard deviation, SIB: unaffected siblings of patients with SSD, X<sup>2</sup>: Chi-square test statistic.

### 3.3. Group differences in [^11^C]UCB-J BP_ND_

Regression analyses revealed that when individuals with SSD were compared to HC, reduced regional [^11^C]UCB-J BP_ND_ was found in the right anterior temporal lobe (lateral part, *p* = 0.04), left subcallosal area (*p* = 0.04), bilateral amygdala (left: *p* = 0.04, right: *p* = 0.03), bilateral parahippocampal and ambient gyrus (left: *p* = 0.02, right: *p* = 0.04), bilateral hippocampus (left: *p* = 0.01, right: *p* = 0.04), inferior frontal gyrus (*p* = 0.03), bilateral superior temporal gyrus (left anterior part: *p* < 0.001, right anterior part: *p* = 0.01; left posterior part: *p* = 0.01, right posterior part: *p* = 0.04), and bilateral insula (left: *p* = 0.01, right: *p* = 0.03). Unaffected siblings of individuals with SSD displayed significantly reduced [^11^C]UCB-J BP_ND_ compared to HC in the left posterior orbital gyrus (*p* = 0.04) and nucleus accumbens (*p* = 0.04), bilateral hippocampus (left: *p* = 0.03, right: *p* = 0.03), bilateral amygdala (left: *p* = 0.03, right: *p* = 0.02), bilateral insula (left: *p* = 0.03, right: *p* = 0.02), the right parahippocampal and ambient gyrus (*p* = 0.02), and bilateral superior temporal gyrus (anterior part, left: *p* = 0.01, right: *p* = 0.01). Figure 1 provides a visual comparison for the mean BP_ND_ per group.

**Fig. 1:**
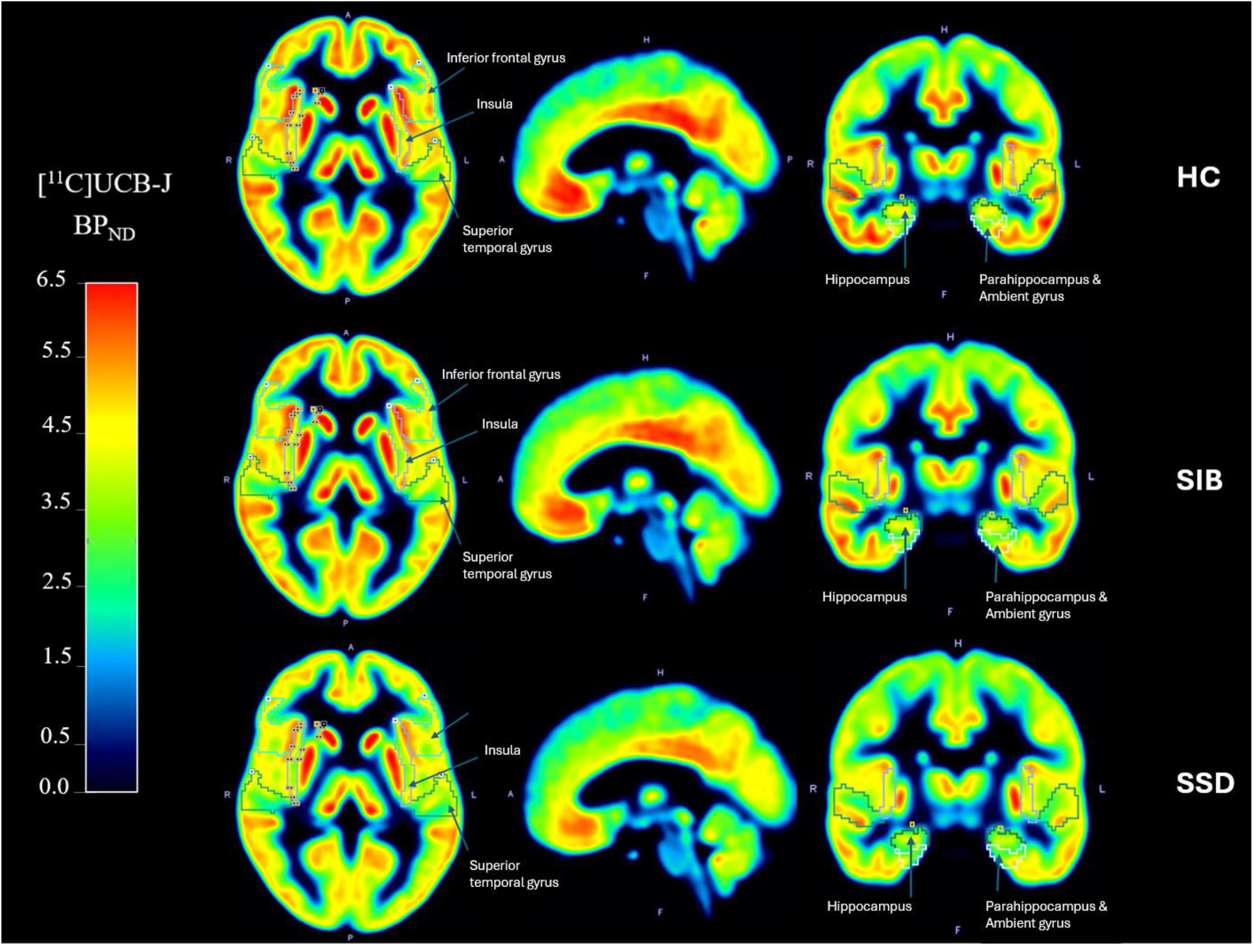
Group means of [^11^C]UCB-J Non-displaceable Binding Potential (BP_ND_) PET-images in Schizophrenia Spectrum Disorder (SSD), unaffected siblings (SIB), and healthy controls (HC). The figure displays the average BP_ND_ PET-images of SSD (*N* = 24) (lower panel), SIB (*N* = 25) (middle panel), and HC (*N* = 26) (upper panel).

Regression analyses including age and years of education as covariates revealed no significant difference between individuals with SSD and unaffected siblings of individuals with SSD (Supplementary table S2.2). These analyses have also been conducted without covariates (see Supplementary table S2.1), yielding similar results.

### 3.4. Relationship of [^11^C]UCB-J BP_ND_ with cognitive and clinical indices

Regressions analyses on verbal memory as main outcome variable and regional [^11^C]UCB-J BP_ND_ and group as predictor variables revealed significant main effects of [^11^C]UCB-J BP_ND_ in the left superior frontal gyrus (*p* = 0.03) and precentral gyrus (*p* = 0.04) and in the right middle frontal gyrus (*p* = 0.04) on verbal memory. For motor speed, significant main effects of bilateral precentral gyri (left: *p* = 0.01; right: *p* = 0.03) as well as cerebellum (left: *p* = 0.003; right: *p* = 0.003) were revealed. For working memory, verbal fluency, executive functioning, attention and information processing speed, none of the assessed ROIs turned out to be significant.

Next, to assess the strength of the relationship between the different cognitive domains and BP_ND_ in ROIs, we performed correlational analyses across groups. These analyses revealed significant relationships between regional [^11^C]UCB-J BP_ND_ and verbal memory performance with the strongest correlation in the left superior frontal gyrus (*r* = 0.39), followed by right middle frontal gyrus (*r* = 0.38), and the bilateral precentral gyrus (left: *r* = 0.35, right: *r* = 0.31) (Supplementary table S3.1). Motor speed and BP_ND_ in both left and right cerebellum were significantly correlated as well (left: *r* = 0.32, right: *r* = 0.33) (Supplementary table S3.2). We assessed the relationship between symptom severity and regional [^11^C]UCB-J BP_ND_. As previously mentioned, the selection of ROIs per PANSS domain was based on previous literature (see supplementary material “*A priori selected ROIs*”). There was no significant relationship between regional [^11^C]UCB-J BP_ND_ and PANSS positive, negative or general scale or the PANSS total score (Supplementary table S4.1 – S4.4).

## 4. Discussion

To elucidate the effects of genetic vulnerability on synaptic density in SSD and its relation to cognition, we assessed synaptic density in 24 individuals with SSD, 25 unaffected siblings of individuals with SSD, and 26 healthy controls, that were similar in sex and age. We confirmed prior findings of widespread bilateral [^11^C]UCB-J BP_ND_ reductions in patients with SSD as compared to HC in the hippocampus, amygdala, and in the frontal and temporal regions. This study is the first to also demonstrate large scale reductions in synaptic density in unaffected siblings of individuals with SSD compared to HC in the same brain regions as in individuals with SSD. Cognitive functioning was significantly lower in the patient group than in HC in overall cognitive functioning and attention and information processing but was unaffected in siblings of individuals with SSD. Compared to unaffected siblings, individuals with SSD showed significant impairment in overall cognitive functioning, verbal memory, and verbal fluency. Despite their adequate cognitive performance, synaptic density of unaffected siblings of individuals with SSD was not different from that of individuals with SSD. This suggests that adequate cognitive performance in unaffected siblings does not preclude affected pruning processes and hints to the use of compensatory strategies or better motivation in siblings. Compensatory mechanisms in the sibling could consist of more efficient functional connectivity [42, 43] or increased white matter efficiency, which can be strengthened by more challenging daily life tasks.

Both individuals with SSD and unaffected siblings showed reduced regional [^11^C]UCB-J BP_ND_ bilaterally in the hippocampus, amygdala, insula and in temporal lobe regions such as the superior temporal gyrus. Individuals with SSD showed significant reductions in the right anterior temporal lobe and left subcallosal area and inferior frontal gyrus. Unaffected siblings did not show reductions in these brain areas. Interestingly, siblings demonstrated additional significant decreases in the left nucleus accumbens and posterior orbital gyrus compared to HC. This finding was not present in the comparison between individuals with SSD and HC.

Husain et al. (2024) reviewed five in-vivo neuroimaging studies on synaptic density in individuals with psychotic disorders, including three in a meta-analysis, and found reductions in frontal, temporal, parietal, occipital areas, amygdala, thalamus, and hippocampus—regions also showing lower synaptic density in our patients and partly also in the unaffected siblings [44]. More recently, Blasco et al. (2025) used [^18^F]SynVesT-1 PET in a small sample of first-episode psychotic patients and individuals at clinical high risk (CHR), finding reduced synaptic density in both groups [25]. They reported prefrontal deficits only in patients and limbic reductions specific to CHR, a finding that parallels our findings in unaffected siblings.

We found only weak correlations between cognitive function and synaptic density across all groups. While we observed significant reductions in [^11^C]UCB-J BP_ND_ in SSD patients and their unaffected siblings, the association between synaptic density and cognitive performance was limited to specific brain regions. In the entire sample, verbal memory performance showed significant positive correlations with BP_ND_ in the left superior frontal gyrus, left precentral gyrus, and right middle frontal gyrus, indicating that higher synaptic density in these regions may support verbal memory functioning. Motor speed was positively correlated with BP_ND_ in the left and right cerebellum, conform the role for cerebellar synaptic integrity in motor performance. The weak correlations we found between synaptic density and cognitive functioning are in agreement with the multi-factorial origin of cognitive impairment in SSD [11, 12]. Also, the demonstration of adequate cognitive performance in siblings, despite widespread deficits in synaptic density illustrates how compensatory mechanisms—such as increased neural efficiency, recruitment of alternative pathways or neurite density [25] — might enable cognitive resilience despite genetic vulnerability and associated synaptic deficits. This is a hopeful finding for researchers who strive to develop interventions to prevent or rescue cognitive functioning in vulnerable groups including those at genetic or clinical high risk.

While previous studies have reported associations between lower regional BP_ND_ and greater positive and negative symptom severity [23, 24, 45], no significant correlations were observed between regional BP_ND_ and PANSS symptom severity (positive, negative, or total scores) in our sample, similar to the findings of Blasco et al. (2025) [25]. While this could be due to a lack of power as our participants did not endure severe symptoms at the time of scanning, it could suggest that synaptic density reductions may primarily reflect a trait marker of genetic vulnerability rather than a direct correlation of clinical symptoms.

### 4.1. Strengths and weaknesses

A key strength of our study is the inclusion of the largest sample size thus far and having groups of SSD patients, unaffected siblings, and healthy controls. Notably, all groups exhibited comparable years of education, which is uncommon in schizophrenia research. The inclusion of siblings without clinical symptoms provided a unique opportunity to examine the impact of genetic vulnerability in the absence of overt disease-related symptoms.

Certain limitations should be acknowledged. While we had detailed information on medication use and duration of disease, we did not have data on cumulative medication use over the years, which would have been valuable. Furthermore, low variation in symptom severity may have obscured potential correlations to synaptic density, as our patient group had relatively low symptom severity. Finally, the use of a reference region might also have limited the sensitivity of the PET scan for determining subtle alterations. For determining subtle alterations, blood sampling should be performed, which enables determination of the individual rate constants and hence might be more sensitive than the reference regio method [46]. Nevertheless, the use of reference regions, however, is considered to be the current standard.

In summary, our findings provide evidence of widespread reductions in synaptic density in individuals with SSD as compared to HC, which were also observed in unaffected siblings. This may suggest that genetic susceptibility to SSD impacts synaptic density. Weak correlations between synaptic density and cognitive functioning were observed in several domains. Although unaffected siblings exhibited substantial reduced synaptic density across multiple brain regions compared to healthy controls, their cognitive performance remained intact. This finding suggests the presence of potential compensatory mechanisms that preserve cognitive function despite underlying synaptic deficits. This encourages research into targeted preventive interventions in young vulnerable groups to prevent cognitive deterioration. Importantly, our results highlight that the absence of cognitive impairments should not be interpreted as an absence of neurobiological deficits in individuals with genetic vulnerability to SSD.

## Data Availability

All data produced in the present study may be based available upon request to the authors and ethical approval.

## Supplementary Information

Supplementary information is available on MP’s website.

## Acknowledgements

The authors wish to express their gratitude to thank Sanne G. Brederoo and Shiral S. Gangadin for their valuable input during the implementation of the study and their thoughtful contributions to the analyses; prof. dr. Anouk van der Hoorn for her radiological guidance and Pernell Willemsen for her unwavering support in practical aspects of the study. We are grateful to all colleagues from the Department of Nuclear Medicine and molecular imaging of the UMCG for their dedicated efforts in tracer production and their professional contributions to the PET-scan procedures. Additionally, we acknowledge all the interns who supported various phases of the study, particular during the data collection and entry. Finally, we would like to thank all participants for showing dedication to participation and investing a significant amount of time in undergoing all study procedures.

## References

1. Schizophrenia. https://www.who.int/news-room/fact-sheets/detail/schizophrenia. Accessed 14 April 2025.

2. Chang WC, Wong CSM, Chen EYH, Lam LCW, Chan WC, Ng RMK, et al. Lifetime prevalence and correlates of schizophrenia-spectrum, affective, and other non-affective psychotic disorders in the Chinese adult population. Schizophr Bull. 2017;43:1280–1290.

3. Habtewold TD, Rodijk LH, Liemburg EJ, Sidorenkov G, Boezen HM, Bruggeman R, et al. A systematic review and narrative synthesis of data-driven studies in schizophrenia symptoms and cognitive deficits. Transl Psychiatry. 2020;10:244.

4. Feber L, Peter NL, Chiocchia V, Schneider-Thoma J, Siafis S, Bighelli I, et al. Antipsychotic Drugs and Cognitive Function. JAMA Psychiatry. 2024. 2024.

5. Galderisi S, Rossi A, Rocca P, Bertolino A, Mucci A, Bucci P, et al. The influence of illness-related variables, personal resources and context-related factors on real-life functioning of people with schizophrenia. World Psychiatry. 2014;13:275–287.

6. Mucci A, Galderisi S, Gibertoni D, Rossi A, Rocca P, Bertolino A, et al. Factors associated with real-life functioning in persons with schizophrenia in a 4-year followup study of the Italian network for research on psychoses. JAMA Psychiatry. 2021;78:550–559.

7. Haddad C, Salameh P, Hallit S, Obeid S, Haddad G, Clément J-P, et al. Cross-cultural adaptation and validation of the Arabic version of the BACS scale (the brief assessment of cognition in schizophrenia) among chronic schizophrenic inpatients. BMC Psychiatry. 2021;21:223.

8. Romanowska S, Best MW, Bowie CR, Depp CA, Patterson TL, Penn DL, et al. Examining the association of life course neurocognitive ability with real-world functioning in schizophrenia-spectrum disorders. Schizophr Res Cogn. 2022;29:100254.

9. Sinkeviciute I, Begemann M, Prikken M, Oranje B, Johnsen E, Lei WU, et al. Efficacy of different types of cognitive enhancers for patients with schizophrenia: a meta-analysis. NPJ Schizophr. 2018;4:22.

10. Howes OD, Dawkins E, Lobo MC, Kaar SJ, Beck K. New drug treatments for schizophrenia: a review of approaches to target circuit dysfunction. Biol Psychiatry. 2024. 2024.

11. Kahn RS, van Rossum IW, Leucht S, McGuire P, Lewis SW, Leboyer M, et al. Amisulpride and olanzapine followed by open-label treatment with clozapine in first-episode schizophrenia and schizophreniform disorder (OPTiMiSE): a three-phase switching study. Lancet Psychiatry. 2018;5:797–807.

12. Faris P, Pischedda D, Palesi F, D’Angelo E. New clues for the role of cerebellum in schizophrenia and the associated cognitive impairment. Front Cell Neurosci. 2024;18:1386583.

13. Feinberg I. Schizophrenia: Caused by a fault in programmed synaptic elimination during adolescence? J Psychiatr Res. 1982;17:319–334.

14. Howes OD, McCutcheon R. Inflammation and the neural diathesis-stress hypothesis of schizophrenia: a reconceptualization. Transl Psychiatry. 2017;7:e1024–e1024.

15. Mana L, Schwartz-Pallejà M, Vila-Vidal M, Deco G. Overview on cognitive impairment in psychotic disorders: From impaired microcircuits to dysconnectivity. Schizophr Res. 2024;269:132–143.

16. Pantelis C, Papadimitriou GN, Papiol S, Parkhomenko E, Pato MT, Paunio T, et al. Biological insights from 108 schizophrenia-associated genetic loci. Nature. 2014;511:421–427.

17. Sekar A, Bialas AR, De Rivera H, Davis A, Hammond TR, Kamitaki N, et al. Schizophrenia risk from complex variation of complement component 4. Nature. 2016;530:177–183.

18. Sellgren CM, Gracias J, Watmuff B, Biag JD, Thanos JM, Whittredge PB, et al. Increased synapse elimination by microglia in schizophrenia patient-derived models of synaptic pruning. Nat Neurosci. 2019;22:374–385.

19. Stanton MM, Modan S, Taylor PM, Hariani HN, Sorokin J, Rash BG, et al. Neuroimmune cortical organoids overexpressing C4A exhibit multiple schizophrenia endophenotypes. BioRxiv. 2023:2021–2023.

20. Finnema SJ, Nabulsi NB, Eid T, Detyniecki K, Lin SF, Chen MK, et al. Imaging synaptic density in the living human brain. Sci Transl Med. 2016;8:1–10.

21. Shanaki Bavarsad M, Spina S, Oehler A, Allen IE, Suemoto CK, Leite REP, et al. Comprehensive mapping of synaptic vesicle protein 2A (SV2A) in health and neurodegenerative diseases: a comparative analysis with synaptophysin and ground truth for PET-imaging interpretation. Acta Neuropathol. 2024;148:58.

22. Onwordi EC, Halff EF, Whitehurst T, Mansur A, Cotel MC, Wells L, et al. Synaptic density marker SV2A is reduced in schizophrenia patients and unaffected by antipsychotics in rats. Nat Commun. 2020;11.

23. Radhakrishnan R, Skosnik PD, Ranganathan M, Naganawa M, Toyonaga T, Finnema S, et al. In vivo evidence of lower synaptic vesicle density in schizophrenia. Mol Psychiatry. 2021;26:7690–7698.

24. Yoon JH, Zhang Z, Mormino E, Davidzon G, Minzenberg MJ, Ballon J, et al. Reductions in synaptic marker SV2A in early-course Schizophrenia. J Psychiatr Res. 2023;161:213–217.

25. Blasco MB, Aji KN, Ramos-Jiménez C, Leppert IR, Tardif CL, Cohen J, et al. Synaptic Density in Early Stages of Psychosis and Clinical High Risk. JAMA Psychiatry. 2025;82:171–180.

26. Tsuang M. Schizophrenia: genes and environment. Biol Psychiatry. 2000;47:210–220.

27. Andreasen NC, Flaum M, Arndt S. The Comprehensive Assessment of Symptoms and History (CASH): an instrument for assessing diagnosis and psychopathology. Arch Gen Psychiatry. 1992;49:615–623.

28. Keefe RSE, Goldberg TE, Harvey PD, Gold JM, Poe MP, Coughenour L. The Brief Assessment of Cognition in Schizophrenia: reliability, sensitivity, and comparison with a standard neurocognitive battery. Schizophr Res. 2004;68:283–297.

29. Stroop JR. Studies of interference in serial verbal reactions. J Exp Psychol. 1935;18:643.

30. Wechsler D. Wechsler adult intelligence scale--. Archives of Clinical Neuropsychology. 1955. 1955.

31. Keefe RSE, Harvey PD, Goldberg TE, Gold JM, Walker TM, Kennel C, et al. Norms and standardization of the Brief Assessment of Cognition in Schizophrenia (BACS). Schizophr Res. 2008;102:108–115.

32. Hammes JGW. De Stroop kleur-woord test. Harcourt Test Publ; 1978.

33. Leucht S, Crippa A, Siafis S, Patel MX, Orsini N, Davis JM. Dose-response meta-analysis of antipsychotic drugs for acute schizophrenia. American Journal of Psychiatry. 2020;177:342–353.

34. Kay SR, Fiszbein A, Opler LA. The Positive and Negative Syndrome Scale (PANSS) for Schizophrenia. Schizophr Bull. 1987;13:261–276.

35. Van Sluis J, De Jong J, Schaar J, Noordzij W, Van Snick P, Dierckx R, et al. Performance characteristics of the digital biograph vision PET/CT system. Journal of Nuclear Medicine. 2019;60:1031–1036.

36. Koole M, van Aalst J, Devrome M, Mertens N, Serdons K, Lacroix B, et al. Quantifying SV2A density and drug occupancy in the human brain using [ 11 C]UCB-J PET imaging and subcortical white matter as reference tissue. Eur J Nucl Med Mol Imaging. 2019;46:396–406.

37. Rossano S, Toyonaga T, Finnema SJ, Naganawa M, Lu Y, Nabulsi N, et al. Assessment of a white matter reference region for 11C-UCB-J PET quantification. Journal of Cerebral Blood Flow and Metabolism. 2020;40:1890–1901.

38. Tuncel H, Boellaard R, Coomans EM, Hollander-Meeuwsen M den, de Vries EFJ, Glaudemans AWJM, et al. Validation and test–retest repeatability performance of parametric methods for [11C] UCB-J PET. EJNMMI Res. 2022;12:3.

39. Peterson RA, Cavanaugh JE. Ordered quantile normalization: a semiparametric transformation built for the cross-validation era. J Appl Stat. 2020. 2020.

40. Peterson RA. Finding optimal normalizing transformations via bestNormalize. 2021. 2021.

41. Cohen J. Statistical power analysis for the behavioral sciences. routledge; 2013.

42. Repovs G, Csernansky JG, Barch DM. Brain network connectivity in individuals with schizophrenia and their siblings. Biol Psychiatry. 2011;69:967–973.

43. Unschuld PG, Buchholz AS, Varvaris M, Van Zijl PCM, Ross CA, Pekar JJ, et al. Prefrontal brain network connectivity indicates degree of both schizophrenia risk and cognitive dysfunction. Schizophr Bull. 2014;40:653–664.

44. Husain MO, Jones B, Arshad U, Ameis SH, Mirfallah G, Schifani C, et al. A systematic review and meta-analysis of neuroimaging studies examining synaptic density in individuals with psychotic spectrum disorders. BMC Psychiatry. 2024;24:460.

45. Onwordi EC, Whitehurst T, Shatalina E, Mansur A, Arumuham A, Osugo M, et al. Synaptic terminal density early in the course of Schizophrenia: an in vivo UCB-J Positron Emission Tomographic Imaging Study of SV2A. Biol Psychiatry. 2024;95:639–646.

46. van der Weijden K. Myelin imaging: past, present, and beyond. 2023. 2023.

