## Supplementary Material for "Synaptic density deficits in patients with a psychotic disorder and unaffected siblings"

#### Synaptic density in patients with a psychotic disorder, their healthy siblings, and healthy participants in relation to cognitions

### Supplementary methods

*Procedure*

Participants were invited for three visits. First, eligibility for participation was assessed during a screening visit. To this end, the Comprehensive Assessment of Symptoms and History (CASH) interview was used to collect demographic information, assess current and past medication intake, and to screen for psychological diseases to determine eligibility for participation (Andreasen et al., 1992). For every individual of the SSD group, the diagnosis was established by the treating psychiatrist as well as validated by means of the CASH-interview. Next, during an ambulatory visit to the study center, participants received an intravenous injection with [11C]UCB-J followed by a PET-scan. Female participants were tested on pregnancy prior to their PET-scan. On the last measurement day, participants underwent a 45min MRI-scan which was followed by 2 hours of cognitive testing.

*Cognitive and Clinical assessments*

The BACS was the neurocognitive assessment battery used in the present study. It consists of the following 7 sub-tests, each assessing a different domain of cognitive functioning:

- *List learning - Verbal memory:* Patients are presented with 15 words and then asked to recall as many as possible, which will be repeated five times. Measure: number of words recalled per trial, in any order.
- *Digit sequencing task - Working memory:* Patients are presented with clusters of numbers of increasing length and are asked to tell them in order from lowest to the highest. Measure: number of correct responses.
- *Token Motor Task - Motor speed:* Patient is given 100 tokens and are asked to place them in a container as quickly as possible. Measure: number of tokens correctly placed into the container.
- *Category instances - Verbal fluency:* Patients are asked to name as many words in a certain category in 60 seconds (supermarket items, tools). Measure: number of unique and appropriate answers per category.
- *Controlled oral word association test - Verbal fluency:* Patients are asked to generate as many words as possible that begin with a given letter in 60 seconds. Measure: number of unique and appropriate answers per category.

- *Symbol coding - Attention and information processing speed:* Timed paper- and-pencil test in which the respondent uses a key to write digits that correspond to nonsense symbols. A sheet with a 9 item key is provided, pairing digit 1–9 with a unique symbol; below are rows of numbers with blank squares beneath. The participant pairs each number with its unique symbol. Measure: number of correct number-symbol pairs completed in 90 seconds.
- *Tower of London - Executive functioning:* Patients are shown two pictures simultaneously with 3 pegs uniquely arranged in each picture. Patients are asked to give the number of times the balls in one picture need to be moved in order to make the arrangements identical on both pictures. There are 20 trials with variable difficulty. Measure: number of correct answers.

*Sample size and power calculation*

Effect sizes reported by Whitfield-Gabrieli et al. (2009) for differences in cognitive performance and functional connectivity between SSD patients and healthy controls as well as between healthy controls and first-degree relatives did not fall below 0.826 (Whitfield-Gabrieli et al., 2009). Matosin et al. (2016) examined synaptic density in post-mortem brain samples of SCZ patients and non-schizophrenic controls and reported an effect size of 1.186 (Matosin et al., 2016). Due to the absence of research on differences in synaptic density between first-degree relatives and healthy controls, we decided to base the presently handled effect size on the most conservative estimation provided by Whitfield-Gabrieli et al. (2009). Although effect sizes of this study are smaller than those reported by Matosin et al. (2016), it is reasonable to still evaluate these as being relatively large, which is the reason for why we chose an effect size of 0.8 for the present analysis. With an effect size of 0.8, a significance level of 0.05, and a desired power of 0.95, a sample size of at least 26 participants per group was required to achieve an actual power of 0.9519 (critical F = 3.0491, numerator df = 3, denominator df = 22).

*A priori ROI selection*

We compared the [$^{11}$C]UCB-J non-displaceable binding potential ($BP_{ND}$) in a set of a priori selected ROIs between groups. To this end, the following ROIs were chosen from a comprehensive range of literature from post-mortem and in-vivo neuroimaging studies which identified brain regions with synaptic protein level abnormalities (Glantz & Lewis, 2000; Osimo et al., 2019) or structural and/or functional alterations (Brugger & Howes, 2017; O'Neill, Mechelli & Bhattacharyya, 2019; Rimol et al., 2010) in SSD: middle frontal gyrus, precentral gyrus, straight gyrus, anterior orbital gyrus, inferior frontal gyrus, superior frontal gyrus, medial orbital gyrus, lateral orbital gyrus, posterior orbital gyrus, subgenual anterior cingulate gyrus, subcallosal area, pre-subgenual anterior cingulate gyrus, hippocampus, amygdala, fusiform gyrus, superior temporal gyrus (anterior and posterior part), anterior temporal lobe (medial and lateral part), middle and inferior temporal gyrus, parahippocampal and ambient gyrus, insula, caudate nucleus, nucleus accumbens, putamen, and thalamus (Glantz & Lewis, 2000; Rimol et al., 2010; Brugger & Howes, 2017; O'Neill, Mechelli & Bhattacharyya, 2019; Osimo et al., 2019).

Furthermore, the association between cognitive functioning and $[^{11}C]$UCB-J $BP_{ND}$ was assessed in *a priori* selected ROIs. These ROIs were chosen based on their association with the particular cognitive domain. That is, verbal memory performance was assessed in the left lateralized medial temporal lobe, referring to the left hippocampus and parahippocampus (Jansen et al., 2009) as well as the bilateral precentral gyrus, left superior frontal gyrus, and right lateralized middle and inferior frontal gyrus (Emch, Bastian & Koch, 2019). Working memory was assessed in the superior frontal cortex (Wager & Smith, 2003) and the right middle frontal gyrus (d'Esposito et al., 1998). Motor speed was examined in the bilateral cerebellum and precentral gyrus (d'Esposito et al., 1998). Verbal fluency was investigated in the cingulate gyrus, right cerebellum (Weiss et al., 2003) and the medial frontal cortex (Cardebat et al., 1996). Attention and information processing speed was assessed in the anterior cingulate cortex (Davis, Hutchison, Lozano, Tasker & Dostrovsky, 2000) and in the left middle frontal gyrus (Turken et al., 2008). Finally, executive functioning was investigated in the left superior parietal cortex (Collette et al., 2005) as well as in the anterior cingulate cortex (Ardila, 2019).

Likewise, the relationship between $[^{11}C]$UCB-J $BP_{ND}$ and the PANSS positive, negative, and general subscales was assessed in *a priori* selected ROIs. The association between $[^{11}C]$UCB-J $BP_{ND}$ and the positive PANSS subscale was assessed in frontal cortices (Padmanabhan et al., 2015; Radhakrishnan et al., 2021), the association with the PANSS negative subscale in the superior frontal gyrus, superior temporal, left anterior cingulate, left insula, and right putamen (Padmanabhan et al., 2015), and the general PANSS subscale in the inferior temporal and pre- and postcentral regions (Menningen et al., 2019).

**Supplementary Table S1: Conversion of categorical education levels to quantitative years of education (YOE).** Procedure handled for the conversion of categorically reported highest completed educational level to quantitative years of education (YOE).

| **Education level in CASH-interview** | | **Years of** |
|---|---|---|
| **Dutch school system** | **American school system** | **education** |
| None | None | 0 |
| Basisschool | Primary school | 6 |
| Lager beroepsonderwijs | Lower vocational education/domestic science school | 10 |
| VMBO/MULO/MAVO | Middle general secondary (vocational) education | 10 |
| Havo | Higher general secondary (vocational) education | 11 |
| VWO, gymnasium | Pre-university education | 12 |
| MBO | Community college/intermediate vocational education | 14 |
| HBO | Higher vocational education | 15 |
| WO/Universiteit | University | 17 |

*CASH: Comprehensive Assessment of Symptoms and History (Andreasen et al., 1992); MAVO: middelbaar algemeen voortgezet onderwijs; MULO: meer uitgebreid lager onderwijs; VMBO: voorbereidend middelbaar beroepsonderwijs; Havo: hogere algemeen voortgezet onderwijs; VWO: voorbereidend wetenschappelijk onderwijs; MBO: middelbaar beroepsonderwijs; HBO: hoger beroepsonderwijs; WO: wetenschappelijk onderwijs.*

### Supplementary results:

#### *Group differences in cognitive performance*

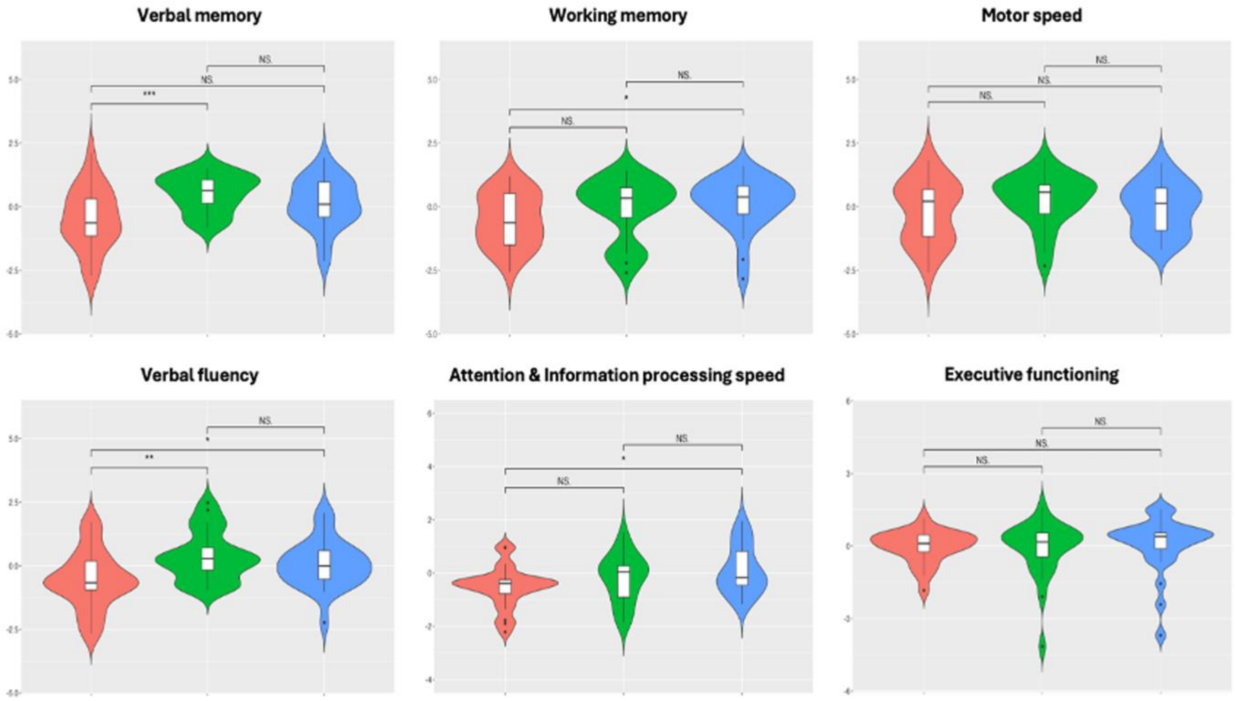


**Figure S1:** ***Violin plots: BACS subtest Z-scores for individuals from the healthy control (HC), unaffected siblings (SIB), and Schizophrenia Spectrum Disorder (SSD) groups***. Y-axis displays the Z-score, X-axis displays the groups (Red: SSD, Green: SIB, Blue: HC). See table 2 for detailed statistics. *BACS: Brief Assessment of Cognition in Schizophrenia.*

### Supplementary results:

#### *Group differences in [$^{11}$C]UCB-J $BP_{ND}$*

**Supplementary Table S2.1.:** ***Results of multiple linear regression analyses to assess differences in [11C]UCB-J $BP_{ND}$ between the groups.*** Statistical analyses of regional [11C]UCB-J $BP_{ND}$ between participants from the SSD, SIB, and HC group and corresponding effect sizes.

| | **SCZ** *(n = 24)* *M/MD (SD/IQR)* | **SIB** *(n = 25)* *M/MD (SD/IQR)* | **HC** *(n = 26)* *M/MD (SD/IQR)* | **Adj. $R^2$** | ***F/H (df)*** | ***p*** | **Cohen's D** | **Post-hoc analyses** |
|---|---|---|---|---|---|---|---|---|
| **Middle Frontal Gyrus, L** | 4.48 (0.61) | 4.76 (0.63) | 4.80 (0.51) | 0.03 | *F*(2, 72) = 2.21 | 0.11 | --- | --- |
| **Middle Frontal Gyrus, R** | 4.41 (0.60) | 4.66 (0.65) | 4.67 (0.46) | 0.01 | *F*(2, 72) = 1.64 | 0.20 | --- | --- |
| **Precentral gyrus, L** | 4.11 (0.58) | 4.24 (0.53) | 4.28 (0.48) | -0.009 | *F*(2, 72) = 0.64 | 0.52 | --- | --- |
| **Precentral gyrus, R** | 4.12 (0.72) | 4.29 (0.56) | 4.41 (0.49) | N/A | *H*(2, 72) = 2.41 | 0.29 | --- | --- |
| **Straight gyrus, L** | 4.59 (0.91) | 4.56 (1.05) | 5.20 (0.89) | N/A | *H*(2, 72) = 9.74 | **0.007**$^{**}$ | 0.677 | *b* |
| **Straight gyrus, R** | 4.44 (1.01) | 4.61 (0.87) | 4.98 (0.88) | N/A | *H*(2, 72) = 9.52 | **0.008**$^{**}$ | 0.572 | *b* |
| **Anterior orbital gyrus, L** | 4.61 (0.74) | 4.76 (0.99) | 5.12 (0.88) | N/A | *H*(2, 72) = 4.68 | 0.09 | --- | --- |
| **Inferior frontal gyrus, L** | 4.46 (0.63) | 4.75 (0.61) | 4.93 (0.51) | 0.07 | *F*(2, 72) = 4.07 | **0.02**$^{*}$ | 0.824 | *b* |
| **Inferior frontal gyrus, R** | 4.52 (0.63) | 4.78 (0.66) | 4.94 (0.47) | 0.05 | *F*(2, 72) = 3.07 | **0.05**$^{*}$ | 0.76 | *b* |
| **Superior frontal gyrus, L** | 4.32 (0.58) | 4.56 (0.65) | 4.66 (0.52) | 0.02 | *F*(2, 72) = 2.11 | 0.12 | --- | --- |
| **Superior frontal gyrus, R** | 4.34 (0.75) | 4.55 (0.74) | 4.76 (0.60) | N/A | *H*(2, 72) = 5.17 | 0.07 | --- | --- |
| **Medial orbital gyrus, L** | 4.51 (0.81) | 4.44 (1.18) | 4.78 (0.55) | N/A | *H*(2, 72) = 5.63 | 0.059 | --- | --- |
| **Medial orbital gyrus, R** | 4.56 (0.87) | 4.65 (1.19) | 5.07 (0.90) | N/A | *H*(2, 72) = 5.31 | 0.07 | --- | --- |
| **Lateral orbital gyrus, L** | 4.24 (0.72) | 4.44 (0.90) | 4.76 (0.59) | N/A | *H*(2, 72) = 5.70 | 0.057 | --- | --- |
| **Posterior orbital gyrus, L** | 4.16 (0.73) | 4.29 (0.99) | 4.73 (0.66) | N/A | *H*(2, 72) = 9.75 | **0.0076**$^{**}$ | 0.821 | *b* |
| **Posterior orbital gyrus, R** | 4.30 (0.66) | 4.43 (1.06) | 4.72 (0.88) | N/A | *H*(2, 72) = 6.82 | **0.03**$^{*}$ | 0.537 | *b* |
| **Subgenual anterior cingulate gyrus, L** | 3.79 (1.19) | 3.81 (0.70) | 4.19 (0.98) | N/A | *H*(2, 72) = 3.26 | 0.19 | --- | --- |
| **Subcallosal area, L** | 3.94 (0.80) | 4.16 (0.61) | 4.81 (0.73) | N/A | *H*(2, 72) = 13.62 | **0.0011**$^{**}$ | 1.138 | *b* |
| **Pre-subgenual anterior cingulate gyrus, L** | 4.85 (1.08) | 4.78 (0.84) | 5.16 (1.33) | N/A | *H*(2, 72) = 2.20 | 0.33 | --- | --- |
| **Pre-subgenual anterior cingulate gyrus, R** | 4.48 (0.95) | 4.41 (0.83) | 4.97 (1.91) | N/A | *H*(2, 72) = 2.67 | 0.26 | --- | --- |
| **Hippocampus, L** | 3.20 (0.44) | 3.35 (0.48) | 3.71 (0.57) | 0.13 | *F*(2, 72) = 6.76 | **0.002**$^{**}$ | *b:* 0.996<br>*c:* 0.682 | *b, c* |
| **Hippocampus, R** | 3.19 (0.42) | 3.29 (0.57) | 3.64 (0.55) | 0.10 | *F*(2, 72) = 5.23 | **0.007**$^{**}$ | 0.915 | *b* |

| | | | | | | | | |
|---|---|---|---|---|---|---|---|---|
| **Amygdala, L** | 3.92 (0.54) | 4 (0.81) | 4.55 (0.80) | 0.10 | *F*(2, 72) = 5.56 | **0.005**** | *b:* 0.916<br>*c:* 0.683 | *b, c* |
| **Amygdala, R** | 3.91 (0.55) | 3.98 (0.89) | 4.62 (0.84) | 0.12 | *F*(2, 72) = 6.48 | **0.002**** | *b:* 0.992<br>*c:* 0.74 | *b, c* |
| **Fusiform gyrus, L** | 4.32 (1.02) | 4.49 (0.95) | 4.93 (1.03) | N/A | *H*(2, 72) = 6.65 | 0.035 | --- | --- |
| **Fusiform gyrus, R** | 3.92 (0.57) | 4.08 (0.67) | 4.44 (0.87) | 0.06 | *F*(2, 72) = 3.42 | **0.038*** | 0.701 | *b* |
| **Superior temporal gyrus (anterior part), L** | 4.10 (0.58) | 4.31 (0.66) | 4.88 (0.78) | 0.17 | *F*(2, 72) = 8.77 | **0.0003***** | *b:* 1.128<br>*c:* 0.788 | *b, c* |
| **Superior temporal gyrus (anterior part), R** | 4.18 (0.61) | 4.32 (0.66) | 4.83 (0.73) | 0.13 | *F*(2, 72) = 6.52 | **0.002**** | *b:* 0.963<br>*c:* 0.732 | *b, c* |
| **Superior temporal gyrus (posterior part), L** | 4.30 (0.56) | 4.56 (0.60) | 4.79 (0.53) | 0.09 | *F*(2, 72) = 4.80 | **0.011*** | 0.90 | *b* |
| **Superior temporal gyrus (posterior part), R** | 4.24 (0.58) | 4.44 (0.59) | 4.63 (0.54) | 0.05 | *F*(2, 72) = 2.98 | 0.05 | --- | --- |
| **Anterior temporal lobe (medial part), L** | 3.88 (0.55) | 3.92 (0.99) | 4.43 (1.04) | 0.05 | *F*(2, 72) = 3.02 | 0.05 | --- | --- |
| **Anterior temporal lobe (medial part), R** | 3.54 (0.61) | 3.68 (0.82) | 4.15 (0.91) | 0.07 | *F*(2, 72) = 4.06 | **0.021*** | 0.781 | *b* |
| **Parahippocampal & ambient gyrus, L** | 3.29 (0.71) | 3.48 (1.09) | 3.77 (1.17) | N/A | *H*(2, 72) = 9.50 | **0.008**** | 0.491 | *b* |
| **Parahippocampal & ambient gyrus, R** | 2.78 (0.40) | 2.86 (0.44) | 3.24 (0.70) | 0.10 | *F*(2, 72) = 5.12 | **0.008**** | *b:* 0.798<br>*c:* 0.647 | *b, c* |
| **Middle & inferior temporal gyrus, L** | 4.56 (0.63) | 4.77 (0.77) | 5.10 (0.82) | 0.05 | *F*(2, 72) = 3.24 | **0.044*** | 0.735 | *b* |
| **Middle & inferior temporal gyrus, Rs** | 4.58 (0.59) | 4.82 (0.71) | 5.12 (0.79) | 0.06 | *F*(2, 72) = 3.57 | **0.033*** | 0.77 | *b* |
| **Anterior temporal lobe (lateral part), L** | 4.43 (0.64) | 4.64 (0.84) | 5.04 (1.09) | 0.05 | *F*(2, 72) = 3.07 | **0.05*** | 0.654 | *b* |
| **Anterior temporal lobe (lateral part), R** | 4.43 (0.65) | 4.63 (0.78) | 5.11 (0.91) | 0.09 | *F*(2, 72) = 4.89 | **0.01*** | 0.854 | *b* |
| **Insula, L** | 4.04 (0.51) | 4.22 (0.55) | 4.54 (0.49) | 0.11 | *F*(2, 72) = 5.83 | **0.004**** | 1.001 | *b* |
| **Insula, R** | 4.09 (0.53) | 4.16 (0.53) | 4.50 (0.49) | 0.08 | *F*(2, 72) = 4.59 | **0.013*** | 0.805 | *b* |
| **Caudate nucleus, L** | 4.72 (0.79) | 4.89 (1.10) | 5.20 (0.75) | 0.02 | *F*(2, 72) = 1.86 | 0.16 | --- | --- |
| **Caudate nucleus, R** | 4.86 (0.88) | 4.99 (0.93) | 5.36 (0.80) | 0.03 | *F*(2, 72) = 2.17 | 0.121 | --- | --- |
| **Nucleus accumbens, L** | 5.36 (0.83) | 5.22 (1.04) | 5.82 (1.17) | N/A | *H*(2, 72) = 7.02 | **0.029*** | 0.541 | *c* |
| **Nucleus accumbens, R** | 4.99 (0.95) | 4.95 (1.22) | 5.35(1.25) | N/A | *H*(2, 72) = 6.49 | **0.038*** | --- | --- |
| **Putamen, L** | 5.48 (0.96) | 5.50 (1.12) | 5.77 (0.79) | N/A | *H*(2, 72) = 4.64 | 0.09 | --- | --- |
| **Putamen, R** | 5.65 (0.95) | 5.64 (0.93) | 5.99 (0.79) | N/A | *H*(2, 72) = 5.06 | 0.07 | --- | --- |
| **Thalamus, L** | 3.94 (0.61) | 4.10 (0.67) | 4.11 (0.46) | -0.010 | *F*(2, 72) = 0.62 | 0.54 | --- | --- |
| **Thalamus, R** | 3.85 (0.58) | 3.96 (0.66) | 3.93 (0.45) | -0.020 | *F*(2, 72) = 0.25 | 0.77 | --- | --- |

*Adj. $R^2$:* adjusted $R^2$, *F*: Regression test statistic, *H*: Kruskal-Wallis test statistic, HC: healthy volunteers, IQR: Interquartile Range, L: left hemisphere, M: Mean, MD: Median, R: right hemisphere, SCZ: schizophrenia patients, SD: Standard deviation, SIB: healthy siblings of patients with SCZ. *a:* SCZ significantly differ from SIB. *b*: SCZ significantly differ from HC. *c*: SIB significantly differ from HC.

#### Supplementary table S2.2 and S2.3:

See table S2.2 and S2.3 (Supplementary Excel-file) for results of multiple linear regression analyses to assess differences in $[^{11}C]$UCB-J $BP_{ND}$ between the groups including age and years of education (YOE) as covariates

### Supplementary results:

#### *Relationship between [$^{11}$C]UCB-J $BP_{ND}$ and Cognitive & clinical indices*

**Supplementary Table S3.1:** ***Correlations between [$^{11}$C]UCB-J $BP_{ND}$ and cognitive performance on the verbal memory subtest of the BACS***. Associations between regional [$^{11}$C]UCB-J $BP_{ND}$ and Z-scores on the verbal memory subtest of the BACS battery.

| | **Composite score** | |
|---|---|---|
| | *r* | *p* |
| Superior Frontal Gyrus, L | 0,394 | **0,001**** |
| Precentral Gyrus, L | 0,358 | **0,002**** |
| Precentral Gyrus, R | 0,310 | **0,007*** |
| Middle Frontal Gyrus, R | 0,383 | **0,001**** |

BACS: Brief Assessment of Cognition in Schizophrenia, L: Left hemisphere, R: Right hemisphere, *r*: Pearsons correlation coefficient, *p*: p-value.

**Supplementary Table S3.2:** ***Correlations between [$^{11}$C]UCB-J $BP_{ND}$ and cognitive performance on the motor speed subtest of the BACS***. Associations between regional [$^{11}$C]UCB-J $BP_{ND}$ and Z-scores on the motor speed subtest of the BACS battery.

| | Composite score | |
|---|---|---|
| | *r* | *p* |
| Cerebellum, L | 0,327 | **0,005**** |
| Cerebellum, L | 0,336 | **0,005**** |
| Precentral Gyrus, L | 0,211 | 0,076 |
| Precentral Gyrus, R | 0,188 | 0,106 |

BACS: Brief Assessment of Cognition in Schizophrenia, L: Left hemisphere, R: Right hemisphere, *r*: Pearsons correlation coefficient, *p*: p-value.

**Supplementary Table S4.1: Correlations between [$^{11}$C]UCB-J $BP_{ND}$ and PANSS scores for the SCZ group.** Associations between regional [$^{11}$C]UCB-J $BP_{ND}$ and scores on PANSS Positive scales.

| **PANSS Positive** | | |
|---|---|---|
| | *r* | *p* |
| Middle Frontal Gyrus, L | -0,264 | 0,236 |
| Middle Frontal Gyrus, R | -0,227 | 0,300 |
| Precentral gyrus, L | -0,246 | 0,264 |
| Precentral gyrus, R | -0,150 | 0,485 |
| Straight gyrus, L | -0,271 | 0,224 |

| | | |
|---|---|---|
| Straight gyrus, R | -0,275 | 0,218 |
| Inferior frontal gyrus, L | -0,255 | 0,248 |
| Inferior frontal gyrus, R | -0,184 | 0,396 |
| Superior frontal gyrus, L | -0,213 | 0,327 |
| Superior frontal gyrus, R | -0,161 | 0,456 |
| Medial orbital gyrus, L | -0,233 | 0,289 |
| Medial orbital gyrus, R | -0,284 | 0,204 |
| Posterior orbital gyrus, L | -0,216 | 0,323 |
| Posterior orbital gyrus, R | -0,255 | 0,248 |

L: Left hemisphere, PANSS: Positive And Negative Syndrome Scale, R: Right hemisphere, *r*: Pearson's correlation coefficient, *p*: p-value.

167

**Supplementary Table S4.2: Correlations between [$^{11}$C]UCB-J $BP_{ND}$ and PANSS scores for the SCZ group.** Associations between regional [$^{11}$C]UCB-J $BP_{ND}$ and scores on PANSS Negative scale.

| **PANSS Negative** | | |
|---|---|---|
| | *r* | *p* |
| Superior Frontal Gyrus, L | 0,033 | 0,940 |
| Superior Frontal Gyrus, R | 0,035 | 0,940 |
| Superior Temporal gyrus (anterior part), L | 0,075 | 0,861 |
| Superior Temporal gyrus (anterior part), R | 0,124 | 0,686 |
| Superior Temporal gyrus (posterior part), L | -0,069 | 0,863 |
| Superior Temporal gyrus (posterior part), R | 0,020 | 0,944 |
| Subgenual Anterior Cingulate gyrus, L | -0,040 | 0,940 |
| Insula, L | 0,015 | 0,944 |

| | | |
|---|---|---|
| Putamen, R | 0,026 | 0,944 |

L: Left hemisphere, PANSS: Positive And Negative Syndrome Scale, R: Right hemisphere, *r*: Pearson's correlation coefficient, *p*: p-value.

**Supplementary Table S4.3: Correlations between [$^{11}$C]UCB-J $BP_{ND}$ and PANSS scores for the SCZ group.** Associations between regional [$^{11}$C]UCB-J $BP_{ND}$ and scores on PANSS General scale.

**PANSS General**

| | *r* | *p* |
|---|---|---|
| Inferior Temporal gyrus, L | -0,014 | 0,947 |
| Inferior Temporal gyrus, R | 0,033 | 0,924 |
| Precentral gyrus, L | -0,141 | 0,630 |
| Precentral gyrus, R | -0,039 | 0,924 |
| Postcentral gyrus, L | -0,217 | 0,404 |
| Postcentral gyrus, R | -0,092 | 0,779 |

L: Left hemisphere, PANSS: Positive And Negative Syndrome Scale, R: Right hemisphere, *r*: Pearson's correlation coefficient, *p*: p-value.

**Supplementary Table S4.4: Correlations between [$^{11}$C]UCB-J $BP_{ND}$ and PANSS scores for the SCZ group.** Associations between regional [$^{11}$C]UCB-J $BP_{ND}$ and scores on PANSS Total scale.

**PANSS Total**

| | *r* | *p* |
|---|---|---|
| Middle Frontal Gyrus, L | -0,157 | 0,490 |
| Middle Frontal Gyrus, R | -0,092 | 0,689 |
| Precentral gyrus, L | -0,172 | 0,447 |
| Precentral gyrus, R | -0,059 | 0,798 |
| Straight gyrus, L | -0,133 | 0,560 |
| Straight gyrus, R | -0,137 | 0,549 |
| Inferior frontal gyrus, L | -0,142 | 0,534 |
| Inferior frontal gyrus, R | -0,075 | 0,746 |
| Superior frontal gyrus, L | -0,093 | 0,687 |
| Superior frontal gyrus, R | -0,049 | 0,828 |
| Medial orbital gyrus, L | -0,088 | 0,702 |
| Medial orbital gyrus, R | -0,138 | 0,547 |
| Posterior orbital gyrus, L | -0,106 | 0,647 |
| Posterior orbital gyrus, R | -0,157 | 0,490 |
| Superior Frontal Gyrus, L | -0,093 | 0,687 |
| Superior Frontal Gyrus, R | -0,049 | 0,828 |
| Superior Temporal gyrus (anterior part), L | -0,029 | 0,897 |
| Superior Temporal gyrus (anterior part), R | -0,031 | 0,891 |
| Superior Temporal gyrus (posterior part), L | -0,134 | 0,558 |
| Superior Temporal gyrus (posterior part), R | -0,031 | 0,891 |
| Subgenual Anterior Cingulate gyrus, L | -0,241 | 0,273 |

| | | |
|---|---|---|
| Insula, L | -0,090 | 0,695 |
| Putamen, R | -0,040 | 0,860 |
| Inferior Temporal gyrus, L | -0,050 | 0,828 |
| Inferior Temporal gyrus, R | -0,005 | 0,981 |
| Precentral gyrus, L | -0,172 | 0,447 |
| Precentral gyrus, R | -0,059 | 0,798 |
| Postcentral gyrus, L | -0,262 | 0,231 |
| Postcentral gyrus, R | -0,120 | 0,601 |

L: Left hemisphere, PANSS: Positive And Negative Syndrome Scale, R: Right hemisphere, *r*: Pearson's correlation coefficient, *p*: p-value.
