## Supplementary Table S2.3 for "Synaptic density deficits in patients with a psychotic disorder and unaffected siblings"

**Supplementary Table S2.3:** ***Results of multiple linear regression analyses to assess diff***
including age and years of education (YoE) as covariates. *Note: regression analyses displ*

| ROI-name | Adjusted R2 | F-statistic | P-value | Intercept: beta-value |
|---|---|---|---|---|
| Middle frontal gyrus, L | 0.21 | 6.06 | 0.00 | 4.17 |
| Middle frontal gyrus, R | 0.19 | 5.47 | 0.00 | 4.24 |
| Precentral gyrus, L | 0.18 | 5.01 | 0.01 | 3.85 |
| Precentral gyrus, R | 0.16 | 4.49 | 0.01 | 3.83 |
| Straight gyrus, L | 0.07 | 2.48 | 0.10 | 3.61 |
| Straight gyrus, R | 0.08 | 2.69 | 0.08 | 3.49 |
| Anterior orbital gyrus, L | 0.06 | 2.11 | 0.15 | 3.23 |
| Inferior frontal gyrus, L | 0.24 | 6.79 | 0.00 | 4.16 |
| Inferior frontal gyrus, R | 0.20 | 5.52 | 0.00 | 4.10 |
| Superior frontal gyrus, L | 0.18 | 5.03 | 0.01 | 4.04 |
| Superior frontal gyrus, R | 0.18 | 5.06 | 0.01 | 4.02 |
| Medial orbital gyrus, L | 0.04 | 1.87 | 0.19 | 3.45 |
| Medial orbital gyrus, R | 0.04 | 1.80 | 0.21 | 3.54 |
| Lateral orbital gyrus, L | 0.08 | 2.72 | 0.08 | 3.03 |
| Posterior orbital gyrus, L | 0.12 | 3.53 | 0.03 | 3.38 |
| Posterior orbital gyrus, R | 0.10 | 3.11 | 0.05 | 3.72 |
| Subgenual anterior cingulate gyrus, L | 0.02 | 1.46 | 0.31 | 3.09 |
| Subcallosal area, L | 0.09 | 2.80 | 0.07 | 3.51 |
| Pre-subgenual anterior cingulate gyrus, L | 0.03 | 1.51 | 0.29 | 3.72 |
| Pre-subgenual anterior cingulate gyrus, R | 0.02 | 1.45 | 0.31 | 3.04 |
| Hippocampus, R | 0.13 | 3.73 | 0.03 | 2.69 |
| Hippocampus, L | 0.15 | 4.34 | 0.01 | 2.74 |
| Amygdala, R | 0.12 | 3.48 | 0.04 | 3.29 |
| Amygdala, L | 0.10 | 3.14 | 0.05 | 3.33 |
| Fusiform gyrus, R | 0.11 | 3.32 | 0.04 | 3.77 |
| Fusiform gyrus, L | 0.04 | 1.70 | 0.24 | 4.04 |
| Superior Temporal gyrus, anterior part, L | 0.20 | 5.76 | 0.00 | 4.08 |
| Superior Temporal gyrus, anterior part, R | 0.16 | 4.63 | 0.01 | 3.93 |
| Superior Temporal gyrus, posterior part, R | 0.20 | 5.76 | 0.00 | 4.19 |
| Superior Temporal gyrus, posterior part, L | 0.24 | 6.79 | 0.00 | 4.34 |
| Anterior Temporal Lobe, medial part, R | 0.09 | 2.76 | 0.08 | 3.02 |
| Anterior Temporal Lobe, medial part, L | 0.06 | 2.23 | 0.13 | 3.20 |
| Parahippocampal & ambient gyrus, R | 0.12 | 3.44 | 0.04 | 2.76 |
| Parahippocampal & ambient gyrus, L | 0.13 | 3.66 | 0.03 | 2.63 |
| Middle & inferior temporal gyrus, R | 0.11 | 3.35 | 0.04 | 4.27 |
| Middle & inferior temporal gyrus, L | 0.10 | 2.97 | 0.06 | 4.33 |
| Anterior Temporal Lobe, lateral part, R | 0.13 | 3.75 | 0.03 | 3.62 |
| Anterior Temporal Lobe, lateral part, L | 0.05 | 1.95 | 0.18 | 4.19 |
| Insula, L | 0.19 | 5.31 | 0.00 | 3.72 |
| Insula, R | 0.17 | 4.76 | 0.01 | 3.75 |
| Caudate nucleus, L | 0.09 | 2.89 | 0.07 | 3.84 |
| Caudate nucleus, R | 0.08 | 2.63 | 0.09 | 4.26 |
| Nucleus accumbens, L | 0.09 | 2.73 | 0.08 | 3.71 |
| Nucleus accumbens, R | 0.05 | 1.94 | 0.18 | 3.95 |
| Putamen, L | 0.08 | 2.62 | 0.09 | 4.73 |
| Putamen, R | 0.07 | 2.48 | 0.10 | 4.78 |

| | | | | |
|---|---|---|---|---|
| Thalamus, L | 0.11 | 3.22 | 0.05 | 3.66 |
| Thalamus, R | 0.09 | 2.81 | 0.07 | 3.70 |

***ferences in [11C]UCB-J BPND between the groups.*** Statistical analyses of regional [11C]UC
*ayed here used the SIB group as reference level.*

| **Intercept: t-value** | **Intercept: p-value** | **SCZ: beta-value** | **SCZ: t-value** | **SCZ: p-value** | **HC: beta-value** | **HC: t-value** | **HC: p-value** |
|---|---|---|---|---|---|---|---|
| 8.76 | 0.00 | -0.12 | -0.73 | 0.57 | 0.18 | 1.15 | 0.34 |
| 8.98 | 0.00 | -0.11 | -0.68 | 0.59 | 0.14 | 0.95 | 0.44 |
| 8.79 | 0.00 | 0.01 | 0.07 | 0.96 | 0.16 | 1.16 | 0.34 |
| 8.67 | 0.00 | -0.02 | -0.14 | 0.92 | 0.18 | 1.28 | 0.29 |
| 4.35 | 0.00 | 0.01 | 0.03 | 0.98 | 0.56 | 2.12 | 0.08 |
| 4.22 | 0.00 | 0.08 | 0.29 | 0.84 | 0.63 | 2.38 | 0.05 |
| 3.83 | 0.00 | 0.20 | 0.72 | 0.57 | 0.52 | 1.92 | 0.11 |
| 8.63 | 0.00 | -0.13 | -0.82 | 0.51 | 0.31 | 1.98 | 0.10 |
| 8.30 | 0.00 | -0.10 | -0.59 | 0.65 | 0.27 | 1.71 | 0.15 |
| 8.29 | 0.00 | -0.09 | -0.56 | 0.67 | 0.21 | 1.37 | 0.26 |
| 8.61 | 0.00 | -0.11 | -0.72 | 0.57 | 0.20 | 1.34 | 0.27 |
| 4.39 | 0.00 | 0.08 | 0.32 | 0.83 | 0.44 | 1.74 | 0.15 |
| 4.30 | 0.00 | 0.07 | 0.24 | 0.88 | 0.44 | 1.69 | 0.16 |
| 4.00 | 0.00 | 0.15 | 0.58 | 0.66 | 0.50 | 2.04 | 0.09 |
| 5.28 | 0.00 | 0.04 | 0.18 | 0.91 | 0.51 | 2.51 | **0.04** |
| 6.09 | 0.00 | 0.01 | 0.06 | 0.97 | 0.45 | 2.28 | 0.06 |
| 3.56 | 0.00 | 0.33 | 1.13 | 0.35 | 0.53 | 1.88 | 0.12 |
| 4.51 | 0.00 | -0.04 | -0.17 | 0.91 | 0.60 | 2.39 | 0.05 |
| 3.81 | 0.00 | 0.34 | 1.04 | 0.40 | 0.59 | 1.90 | 0.11 |
| 2.84 | 0.02 | 0.34 | 0.96 | 0.43 | 0.62 | 1.79 | 0.13 |
| 5.76 | 0.00 | -0.01 | -0.07 | 0.96 | 0.39 | 2.60 | **0.03** |
| 6.10 | 0.00 | -0.06 | -0.40 | 0.77 | 0.39 | 2.70 | **0.03** |
| 4.65 | 0.00 | 0.01 | 0.03 | 0.98 | 0.65 | 2.86 | **0.02** |
| 5.02 | 0.00 | 0.00 | 0.02 | 0.99 | 0.57 | 2.67 | **0.03** |
| 5.96 | 0.00 | -0.05 | -0.22 | 0.89 | 0.46 | 2.29 | 0.06 |
| 5.18 | 0.00 | -0.13 | -0.48 | 0.72 | 0.33 | 1.30 | 0.28 |
| 6.74 | 0.00 | -0.12 | -0.60 | 0.65 | 0.66 | 3.38 | **0.01** |
| 6.61 | 0.00 | -0.04 | -0.19 | 0.90 | 0.59 | 3.10 | **0.01** |
| 8.89 | 0.00 | -0.07 | -0.47 | 0.73 | 0.32 | 2.14 | 0.08 |
| 9.28 | 0.00 | -0.14 | -0.91 | 0.46 | 0.36 | 2.40 | 0.05 |
| 4.24 | 0.00 | -0.03 | -0.12 | 0.94 | 0.52 | 2.29 | 0.06 |
| 3.99 | 0.00 | 0.08 | 0.29 | 0.84 | 0.57 | 2.23 | 0.07 |
| 5.72 | 0.00 | -0.02 | -0.14 | 0.92 | 0.44 | 2.82 | **0.02** |
| 4.39 | 0.00 | -0.08 | -0.42 | 0.76 | 0.42 | 2.18 | 0.07 |
| 6.86 | 0.00 | -0.11 | -0.53 | 0.69 | 0.38 | 1.93 | 0.11 |
| 6.54 | 0.00 | -0.08 | -0.38 | 0.79 | 0.43 | 2.00 | 0.10 |
| 5.18 | 0.00 | -0.05 | -0.20 | 0.90 | 0.54 | 2.40 | 0.05 |
| 5.24 | 0.00 | -0.11 | -0.43 | 0.76 | 0.46 | 1.79 | 0.13 |
| 8.27 | 0.00 | -0.07 | -0.47 | 0.73 | 0.39 | 2.70 | **0.03** |
| 8.38 | 0.00 | 0.03 | 0.23 | 0.89 | 0.42 | 2.95 | **0.02** |
| 4.94 | 0.00 | 0.02 | 0.08 | 0.96 | 0.42 | 1.67 | 0.16 |
| 5.58 | 0.00 | 0.04 | 0.15 | 0.92 | 0.48 | 1.95 | 0.11 |
| 3.95 | 0.00 | 0.24 | 0.78 | 0.54 | 0.75 | 2.50 | **0.04** |
| 4.49 | 0.00 | 0.10 | 0.35 | 0.81 | 0.61 | 2.17 | 0.08 |
| 7.55 | 0.00 | 0.06 | 0.31 | 0.83 | 0.41 | 2.03 | 0.09 |
| 6.77 | 0.00 | 0.16 | 0.66 | 0.60 | 0.53 | 2.36 | 0.05 |

| | | | | | | | |
|---|---|---|---|---|---|---|---|
| 7.35 | 0.00 | -0.03 | -0.15 | 0.92 | 0.12 | 0.76 | 0.55 |
| 7.61 | 0.00 | 0.00 | -0.01 | 0.99 | 0.08 | 0.51 | 0.70 |

CB-J BPND between participants from the SCZ, SIB, and HC group,

| **Age: beta-value** | **Age: t-value** | **Age: p-value** | **YoE: beta-value** | **YoE: t-value** | **YoE: p-value** |
|---|---|---|---|---|---|
| -0.02 | -3.47 | **0.00** | 0.09 | 3.05 | **0.01** |
| -0.02 | -3.57 | **0.00** | 0.08 | 2.74 | **0.03** |
| -0.02 | -3.63 | **0.00** | 0.07 | 2.77 | **0.02** |
| -0.02 | -3.26 | **0.01** | 0.07 | 2.60 | **0.03** |
| -0.01 | -0.68 | 0.59 | 0.08 | 1.64 | 0.17 |
| -0.01 | -0.68 | 0.59 | 0.09 | 1.69 | 0.16 |
| 0.00 | -0.48 | 0.72 | 0.10 | 2.02 | 0.09 |
| -0.02 | -3.30 | **0.01** | 0.09 | 2.95 | **0.02** |
| -0.02 | -2.89 | **0.02** | 0.09 | 2.91 | **0.02** |
| -0.02 | -3.13 | **0.01** | 0.08 | 2.72 | **0.03** |
| -0.02 | -2.99 | **0.02** | 0.08 | 2.71 | **0.03** |
| -0.01 | -0.91 | 0.46 | 0.09 | 1.78 | 0.14 |
| -0.01 | -0.69 | 0.59 | 0.09 | 1.73 | 0.15 |
| -0.01 | -1.06 | 0.39 | 0.11 | 2.33 | 0.06 |
| -0.01 | -1.32 | 0.27 | 0.08 | 2.10 | 0.08 |
| -0.01 | -1.53 | 0.20 | 0.07 | 1.91 | 0.11 |
| -0.01 | -1.17 | 0.33 | 0.08 | 1.47 | 0.22 |
| -0.01 | -1.00 | 0.41 | 0.06 | 1.29 | 0.29 |
| -0.01 | -0.99 | 0.41 | 0.09 | 1.56 | 0.19 |
| 0.00 | -0.20 | 0.90 | 0.10 | 1.55 | 0.19 |
| -0.01 | -1.00 | 0.41 | 0.05 | 1.89 | 0.11 |
| 0.00 | -0.72 | 0.57 | 0.05 | 1.82 | 0.13 |
| 0.00 | 0.03 | 0.98 | 0.04 | 1.03 | 0.40 |
| 0.00 | -0.28 | 0.85 | 0.05 | 1.23 | 0.31 |
| -0.01 | -2.09 | 0.08 | 0.06 | 1.56 | 0.19 |
| -0.01 | -1.26 | 0.30 | 0.06 | 1.29 | 0.29 |
| -0.01 | -1.89 | 0.12 | 0.05 | 1.36 | 0.26 |
| -0.01 | -1.76 | 0.14 | 0.06 | 1.58 | 0.19 |
| -0.02 | -3.53 | **0.00** | 0.07 | 2.33 | 0.06 |
| -0.02 | -3.53 | **0.00** | 0.06 | 2.27 | 0.06 |
| -0.01 | -0.96 | 0.43 | 0.06 | 1.49 | 0.21 |
| -0.01 | -0.99 | 0.41 | 0.07 | 1.47 | 0.22 |
| -0.01 | -1.63 | 0.17 | 0.03 | 1.03 | 0.40 |
| -0.01 | -1.04 | 0.40 | 0.07 | 2.05 | 0.09 |
| -0.01 | -1.79 | 0.13 | 0.07 | 1.85 | 0.12 |
| -0.01 | -1.80 | 0.13 | 0.06 | 1.61 | 0.18 |
| -0.01 | -1.03 | 0.40 | 0.09 | 2.07 | 0.09 |
| -0.01 | -0.90 | 0.46 | 0.05 | 1.05 | 0.39 |
| -0.01 | -2.13 | 0.08 | 0.06 | 2.27 | 0.06 |
| -0.01 | -2.34 | 0.06 | 0.06 | 2.17 | 0.08 |
| -0.02 | -1.75 | 0.14 | 0.11 | 2.33 | 0.06 |
| -0.01 | -1.77 | 0.14 | 0.09 | 1.90 | 0.11 |
| 0.00 | 0.18 | 0.90 | 0.09 | 1.63 | 0.17 |
| 0.00 | -0.33 | 0.83 | 0.07 | 1.24 | 0.31 |
| -0.01 | -1.35 | 0.26 | 0.08 | 2.05 | 0.09 |
| -0.01 | -1.13 | 0.35 | 0.08 | 1.80 | 0.13 |

| | | | | | |
|---|---|---|---|---|---|
| -0.02 | -2.76 | **0.02** | 0.07 | 2.32 | 0.06 |
| -0.02 | -2.85 | **0.02** | 0.06 | 1.98 | 0.10 |
